# Long-Term Acceptability of Hygiene, Face Covering, and Social Distancing Interventions to Prevent Exacerbations in people living with Airways Diseases

**DOI:** 10.1101/2021.04.09.21255189

**Authors:** John R Hurst, Andrew Cumella, Camila Nagoda Niklewicz, Keir EJ Philip, Victoria Singh, Nicholas S Hopkinson

## Abstract

**Introduction:** There has been a substantial reduction in admissions to hospital with exacerbations of airways diseases during the COVID-19 pandemic, likely because measures introduced to prevent the spread of SARS-CoV-2 also reduced transmission of other respiratory viruses. The acceptability to patients of continuing such interventions beyond the pandemic as a measure to prevent exacerbations is not known.

**Method:** An online survey of people living with respiratory disease was created by the Asthma UK – British Lung Foundation Partnership. People were asked what infection control measures they expected to continue themselves, and what they thought should be policy for the population more generally in the future, once the COVID-19 pandemic had subsided.

**Results:** 4442 people completed the survey: 3627 with asthma, 258 with bronchiectasis and 557 with COPD. Regarding personal behaviour, 79.5% would continue increased handwashing, 68.6% social distancing indoors, 46.9% would continue to wear a face covering in indoor public places (45.7% on public transport), and 59.3% would avoid friends and family who were unwell with a respiratory infection. 45.6% wanted healthcare professionals to continue wearing a mask when seeing patients. 60.7% thought that face coverings should continue to be worn by everyone in indoor public spaces during the ‘flu season. Women and older people were, in general, more cautious.

**Conclusion:** People living with airways diseases are supportive of infection control measures to reduce the risk of exacerbations and such measures should be considered for inclusion in guidelines. Further research to refine understanding of the most effective approaches is needed.

## Introduction

Chronic airway diseases comprising asthma, bronchiectasis and chronic obstructive pulmonary disease (COPD) are very common [1-3]. People living with these conditions are susceptible to recurrent deteriorations in their respiratory health called ‘exacerbations’ which cause much of the morbidity, mortality, and health-care costs [1-3]. Patients with COPD, for example, rate exacerbations as the most disruptive aspect of their disease [4]. Most exacerbations are caused by acquisition of airway infection, particularly with respiratory viruses including coronaviruses and rhinoviruses [5-7].

During the COVID-19 pandemic, a range of measures to reduce the transmission of SARS-CoV-2 were introduced including encouraging hand hygiene, social (physical) distancing, and the wearing of face coverings. People living with chronic airway diseases were thought, and subsequently shown to be at increased risk of poor outcomes with SARS-CoV-2, the virus that causes COVID-19 [8, 9]. Such individuals, perceived to be particularly vulnerable, chose or were advised to take additional precautions including ‘shielding’ (maximal reduction of face-to-face social contact [10]) during the pandemic. Such recommendations varied within and between countries, and over time.

The pandemic was associated with a dramatic fall in cases of influenza [11] and multiple studies have reported a substantial reduction in hospitalisations due to asthma and COPD exacerbations, temporally associated with the COVID-19 pandemic including introduction of respiratory virus infection control measures [12-20], likely because vulnerable people avoiding SARS-CoV-2 were also avoiding other respiratory viruses that are common causes of exacerbations [21].

Current clinical guidelines in chronic airway diseases such as the Global Initiative for Asthma (GINA [1]), Global Initiative for Chronic Obstructive Lung Disease (GOLD [3]) and the European Respiratory Society (ERS) guidelines on bronchiectasis [2] focus on exacerbation prevention but do not specifically recommend respiratory virus infection control measures. This is despite a Cochrane review first published in 2011 suggesting that such interventions can be effective [22].

The COVID-19 pandemic has demonstrated that individuals both with and without long term health conditions are prepared to accept measures to reduce the transmission of infection. This can be driven both by a desire to protect the self, or to protect others, and will reflect national guidance or legislation as well as personal beliefs about what is effective. Understanding the attitudes of people living with respiratory disease is an important first step in developing policy and guidelines in this area.

## Methods

An online survey of people with respiratory disease was created by the Asthma UK – British Lung Foundation (AUK-BLF) partnership, the largest UK respiratory disease charity, as part of ongoing work to evaluate the impact of the COVID-19 pandemic and guide responses to it. The survey was distributed and promoted via AUK-BLF social media and mailing lists and open for responses between the 3^rd^ and 10^th^ of March 2021. Data collection was via Typeform. Data were collected on demographics and self-reported respiratory diagnosis. As part of the survey, respondents were asked their views about measures to reduce respiratory virus transmission. These questions were deliberately framed to cover a future period after the COVID-19 pandemic was over, based on the possibility that such measures could reduce the risk of exacerbations of lung disease. We used the following wording: “*There is some evidence that the measures to control spread of COVID-19 during the pandemic have also reduced the spread of other viruses (such as colds and ‘flu) that can cause asthma attacks / exacerbations of lung disease. Thinking about the future (after the lockdown has been eased and most people have been vaccinated against COVID-19), which of the following…*” (see Table 1). The survey asked what people expected themselves to be doing to reduce exacerbation risk one year from now, as well as what recommendations should be in force for the wider population both “all the time” and “during the ‘flu season”.

**Table 1:** Acceptability of respiratory virus infection control interventions in people living with asthma (n=3627), bronchiectasis (n=258) and COPD (n=557). Data are % answering yes (total n=4442). Green ≥66%, Red ≤33%. Shading indicates ≥10% points difference across diseases.

|  | Thinking about the future (after the lockdown has been eased and most people have been vaccinated against COVID-19), would you *continue to do yourself, even if it wasn't policy...* | Thinking about the future (after the lockdown has been eased and most people have been vaccinated against COVID-19), which of the following do you think should carry on as policies *for everyone at all times?* | Thinking about the future (after the lockdown has been eased and most people have been vaccinated against COVID-19), which of the following do you think should carry on as policies *for everyone during the 'flu season?* |
| --- | --- | --- | --- |
| Wearing a face covering in indoor public places | Overall 46.9%<br>Asthma 46.0%<br>Bronchiectasis 56.2%<br>COPD 48.5% | Overall 44.9%<br>Asthma 43.5%<br>Bronchiectasis 54.3%<br>COPD 50.3% | Overall 60.7%<br>Asthma 60.0%<br>Bronchiectasis 70.5%<br>COPD 61.0% |
| Wearing a face covering on public transport | Overall 45.7%<br>Asthma 45.6%<br>Bronchiectasis 51.6%<br>COPD 43.4% | Overall 50.8%<br>Asthma 50.1%<br>Bronchiectasis 55.8%<br>COPD 53.0% | Overall 59.5%<br>Asthma 59.5%<br>Bronchiectasis 63.6%<br>COPD 57.8% |
| Wearing a face covering outdoors | Overall 11.6%<br>Asthma 11.5%<br>Bronchiectasis 12.4%<br>COPD 11.8% | Overall 10.9%<br>Asthma 10.6%<br>Bronchiectasis 11.2%<br>COPD 13.1% | Overall 12.9%<br>Asthma 12.5%<br>Bronchiectasis 14.7%<br>COPD 15.3% |
| Hand sanitiser being widely available to clean hands | Not applicable | Overall 79.7%<br>Asthma 80.9%<br>Bronchiectasis 81.09%<br>COPD 71.3% | Overall 81.7%<br>Asthma 83.1%<br>Bronchiectasis 81.4%<br>COPD 72.3% |
| Washing hands more often | Overall 79.5%<br>Asthma 80.7%<br>Bronchiectasis 81.0%<br>COPD 70.6% | Overall 73.7%<br>Asthma 74.7%<br>Bronchiectasis 76.7%<br>COPD 65.7% | Overall 75.9%<br>Asthma 76.8%<br>Bronchiectasis 79.1%<br>COPD 68.4% |
| Keeping more of a distance from others when in an indoor public space | Overall 68.6%<br>Asthma 67.8%<br>Bronchiectasis 73.6%<br>COPD 70.9% | Overall 52.5%<br>Asthma 50.8%<br>Bronchiectasis 58.89%<br>COPD 60.3% | Overall 62.9%<br>A 61.4%<br>Bronchiectasis 70.5%<br>COPD 68.6% |
| Keeping more of a distance from others when in an outdoor public space | Overall 38.0%<br>Asthma 37.8%<br>Bronchiectasis 38.0%<br>COPD 39.5% | Not asked | Not asked |
| Avoiding busy public spaces | Overall 60.0%<br>Asthma 58.9%<br>Bronchiectasis 71.7%<br>COPD 61.8% | Not asked | Not asked |
| Avoid seeing friends or family if they are unwell with a cold or 'flu | Overall 59.3%<br>Asthma 59.9%<br>Bronchiectasis 70.2%<br>COPD 50.8% | Not asked | Not asked |
| Healthcare staff to wear masks when seeing patients | Not applicable | Overall 45.6%<br>Asthma 45.5%<br>Bronchiectasis 50.8%<br>COPD 43.6% | Overall 51.5%<br>Asthma 51.6%<br>Bronchiectasis 56.2%<br>COPD 48.7% |

Data were collated in Excel and analysed in SPSS (version 25). Proportions across groups were compared with Chi Square analysis. p≤0.05 was taken to indicate statistical significance. A two thirds or more majority was arbitrarily and *a priori* chosen as indicating general support for a measure, whilst a one third or fewer proportion was taken as indicating the absence of significant support. A difference of more than ten percentage points between groups was arbitrarily and *a priori* chosen to indicate a potentially meaningful difference.

Ethical approval for this study was obtained from the Imperial College Research Governance and Integrity Team (RGIT) (ICREC Ref: 20IC6625). Survey respondents consented to use of their responses for analysis and publication.

## Results

The survey was completed by 4442 people living with airways diseases, including 3627 (81.7%)with asthma, 258 (5.8%) with bronchiectasis and 557 (12.5%) with COPD. The commonest age categories were 50-59 (1165 respondents) and 60-69 years (1200 respondents) with 1218 younger and 802 older participants. 3381 (76%) participants were female, 1013 (23%) were male, and six (0.02%) responded ‘other’ or did not leave a response. 4260 (96%) self-reported their ethnic group as ‘white’.

Responses on the longer-term acceptability of respiratory virus infection control measures by people living with chronic airways diseases, in relation to personal choice, policy and regulation, are presented in Table 1. Given the high proportion of respondents with asthma, results are also presented separately for asthma, bronchiectasis and COPD.

The main finding is that a substantial proportion of respondents expected that they would continue to take some steps to reduce their future risk of exacerbation. 79.5% would continue increased handwashing and 68.6% would continue social distancing indoors. 46.9% would continue to wear a face mask in indoor public places (45.7% on public transport), as well as avoiding friends and family who are unwell with a cold or ‘flu (59.3%). There was little support for wearing face coverings or social distancing outdoors (11.6% and 38.0% respectively).

Regarding policy for people more generally, responses were similar between what the population should be asked to do “at all times” compared to “during the ‘flu season”, except for wearing a face covering indoors (44.9% vs 60.7%) and social distancing indoors (52.5% vs 62.9%). Around half of this population with long term respiratory conditions would want healthcare professionals to continue the current COVID-19 pandemic practice of wearing a mask when seeing patients.

Women were, in general, more likely to plan to continue measures to reduce viral transmission compared to men, maintaining indoor social distancing (71.6% vs. 58.6%, p<0.001), avoiding busy public spaces (62.6% vs. 51.6%, p<0.001) and avoiding friends and family who are unwell (63.5% vs. 45.1%, p<0.001).

Given the lower proportion of people with COPD and bronchiectasis, and the potential for them to differ from people with asthma, we examined the effect of age on respiratory virus infection control measures in people with asthma alone. The data are reported in Table 2. There were statistically significant differences together with differences of more than 10% points between groups by age in willingness to wear a face covering indoors and on public transport, social distancing in an indoor public space, avoiding busy public spaces and avoiding friends and family who are unwell (Figure 1). In general, older people (except the oldest old) were more cautious, with the exception of not visiting friends and family who are unwell where younger people were more cautious.

**Table 2:**
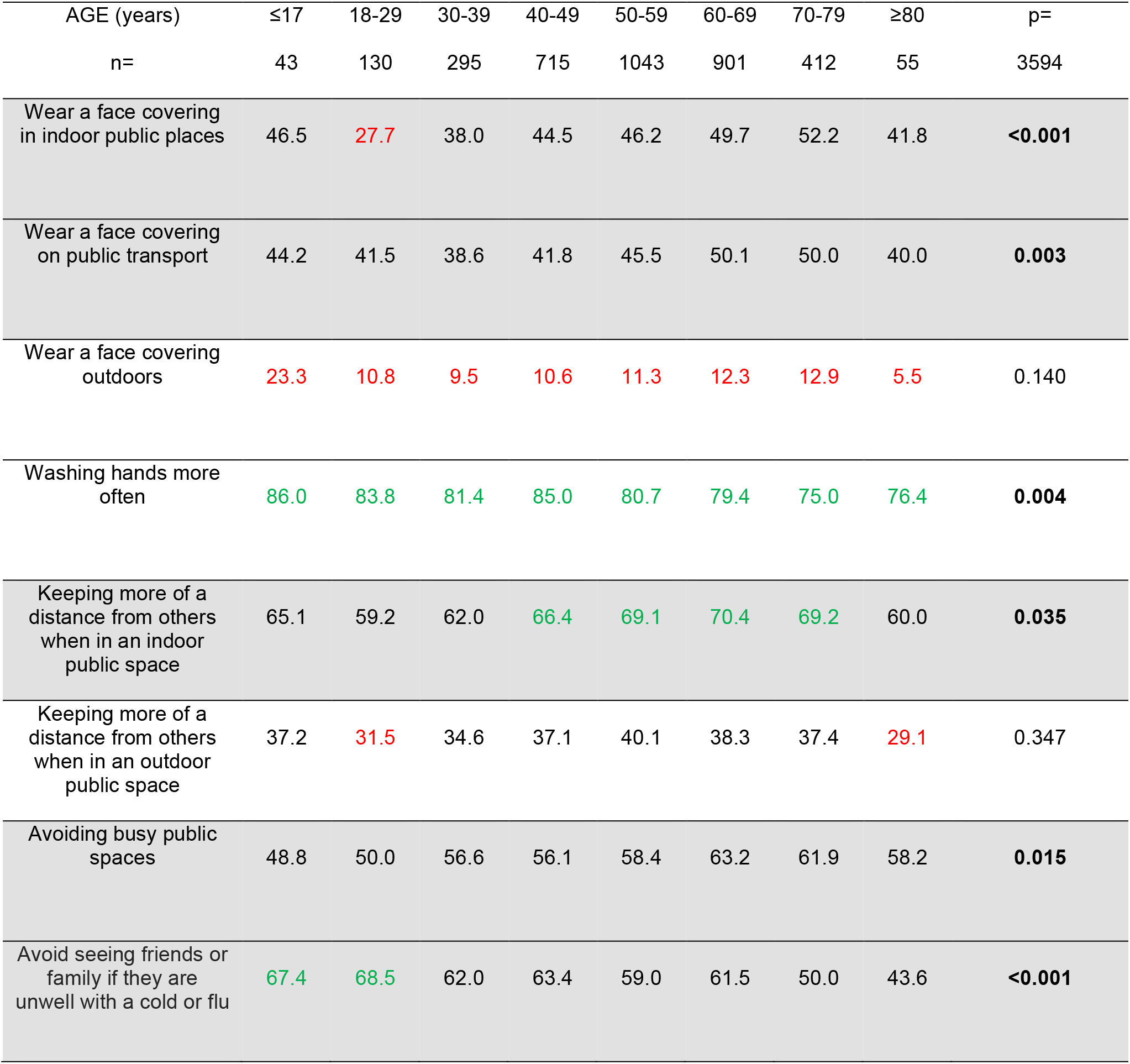
Respiratory virus infection control measures that people living with asthma anticipate they will continue doing one year from now (n=3627), by age. Data are % answering yes. Green ≥66%, Red ≤33%. Shading indicates ≥10% points difference across age categories. p= Chi Square.

**Figure 1:**
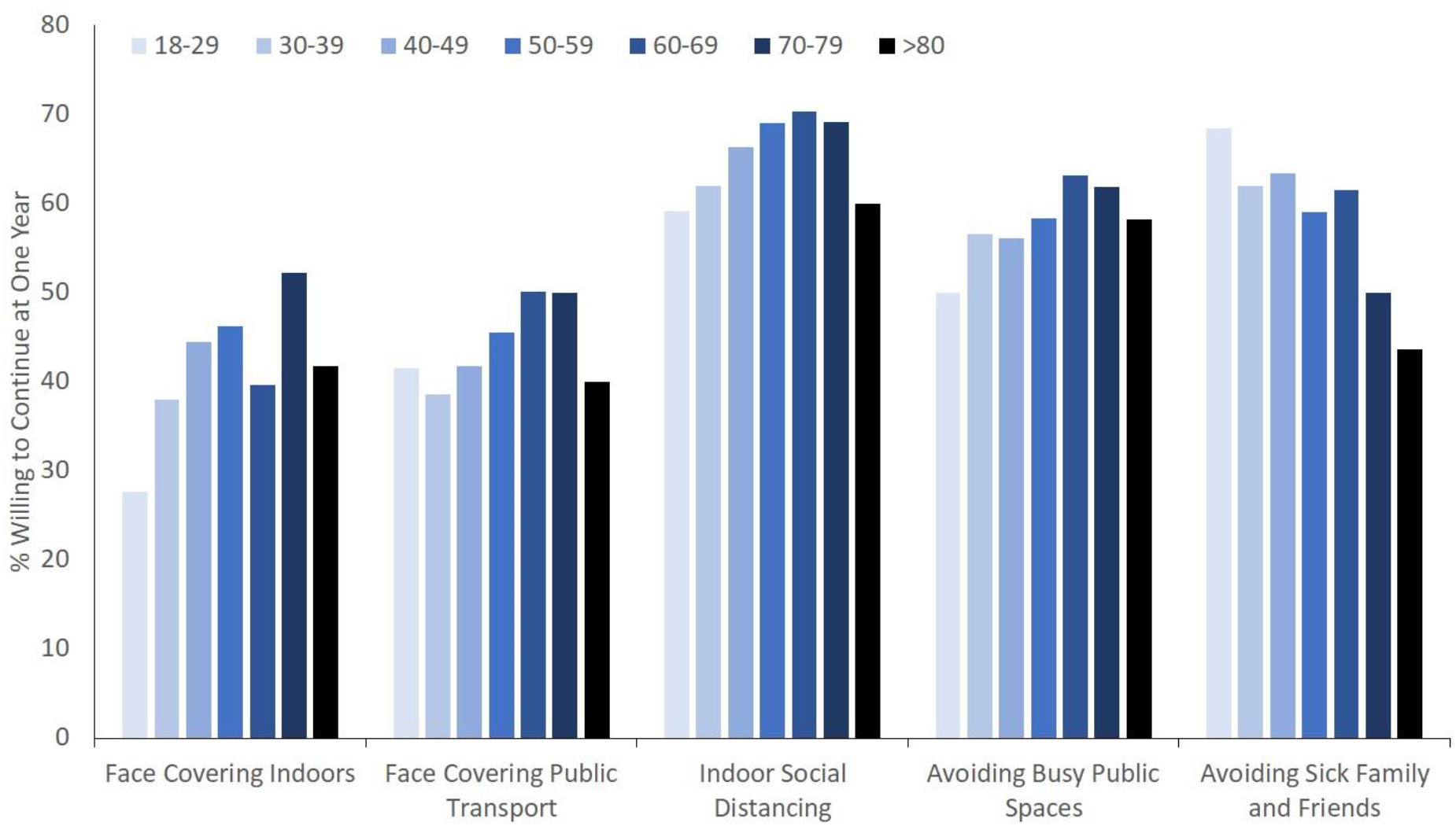
The acceptability of respiratory virus infection control interventions in adults living with asthma, by age (in years). n=3627.

## Discussion

The results of this UK study of 4442 people living with asthma, bronchiectasis and COPD suggests that many intend to continue to adopt respiratory virus infection control measures, first employed during the COVID-19 pandemic, to reduce their future risk of exacerbations. There was also support for the adoption of such measures in the general population during the ‘flu season. Given the evidence base supporting the effectiveness of these measures to reduce exacerbations, consideration should be given to including such measures in guidelines for airways diseases.

The basic premise of our survey was that widespread, continued implementation of pandemic infection control measures was able to reduce the transmission of other viruses which in turn reduced exacerbations of airways diseases in both community and hospital settings [12-20] (noting that other circumstances of the pandemic such as cleaner air may also have contributed). Importantly, the pattern of residual disease severity does not support the counter-hypothesis that this reduction is due to patients avoiding healthcare during the pandemic. The reduction in hospitalisation is of greater magnitude than ever previously achieved with optimisation of non-pharmacological and pharmacological care [1-3]. For example, Tan *et al* reported a 50% reduction in hospitalised COPD [14] (and asthma [17]) exacerbations in Singapore, and a reduction in the proportion of residual exacerbations associated with respiratory viruses. In a hospital in Hong Kong, admissions for COPD exacerbations decreased by 44% during the first three months of 2020 compared with the previous year [13]. In a UK hospital, total COPD and asthma exacerbation admissions in 2020 were 44% and 40% respectively of those in 2019 [16], without an increase in severity. COPD exacerbation admissions at a German hospital fell by 42% during the pandemic, a reduction greater than that seen in myocardial infarction or stroke [12]. We are not aware of any data in relation to bronchiectasis. Although one small study in the UK suggested an increase in community exacerbations [15], robust population data in asthma suggests a reduction in both community [20] and hospital treated events [18] without an increase in asthma deaths, arguing against people inappropriately avoiding hospital care. In the UK, there has also been a striking reduction in community prescriptions for oral corticosteroids [23]. Thus, whilst affecting care delivery and health, including the mental health [24, 25], of people living with chronic airways diseases, the COVID-19 pandemic and ensuing restrictions have demonstrated the potential for dramatically reducing the future burden of exacerbations of airways diseases.

Exacerbations of airways diseases cause a major burden to patients and health services [26] and preventing exacerbations is a cornerstone of respiratory care. Currently, ‘infection control’ measures against micro-organisms associated with exacerbations are not part of routine guidelines including those for asthma (GINA [1]), bronchiectasis (ERS [2]) and COPD (GOLD [3]). This is despite guidelines highlighting the importance of patient education and training in “essential skills” such as inhaler technique and self-management. Primary prevention of exacerbations focuses on optimisation of medical treatment and smoking cessation. Interventions to reduce the acquisition of respiratory viruses are a notable omission. A Cochrane review of physical interventions to reduce the spread of respiratory viruses concluded that simple interventions had the potential to reduce transmission of respiratory viruses while acknowledging that routine, long-term use would be challenging outside the context of an epidemic [22]. However, the profound reduction in exacerbations that has been seen during the pandemic period and the attitudes elicited by our survey suggests that guideline recommendations around respiratory virus infection control measures for people living with chronic respiratory diseases should be revisited.

The questions we asked people living with airways diseases were framed around the potential benefit of continuing respiratory virus infection control measures in the light of evidence that they may be effective at preventing exacerbations. We have shown that there is wide support for continuing measures such as general hand hygiene and the continuing provision of hand sanitiser in public spaces. Whilst over half the respondents thought there should be a recommendation for everyone to wear a mask in indoor public spaces (especially during the (northern hemisphere) Winter), fewer (47%) would personally continue to do this in the absence of such a recommendation. There was more support for continuing ‘social’ (physical) distancing, avoidance of busy public spaces, and restricting visits with friends and family who are unwell. We observed some differences between the major airways diseases (participants with bronchiectasis were the most supportive of continuing infection control measures), between male versus female respondents, and by age. In general, older people (except the oldest old) and women were more willing to adopt a more cautious approach and were more likely to choose to continue with preventative measures even where these were not mandated. The exception to this was willingness to visit family and friends who are unwell, where younger people were most cautious.

More frequent hand washing would be the simplest measure to encourage as it is already part of widely accepted social behaviour, meaning there would be little behaviour change involved. A randomised control trial in primary care in the UK has demonstrated that a web-based hand-washing intervention maximised handwashing behaviours and improved infection rates over three Winters [27]. If it is measures other than hand hygiene that are the most effective at reducing transmission, further work will be required to support people living with airways diseases in making additional behaviour changes. Factors determining compliance with advice in the general population have recently been described and were most closely related to confidence in the source of advice [28]. Pulmonary Rehabilitation could provide a forum in which to disseminate such messages to people living with airways diseases. Other interventions could include reducing presenteesism, where people go to work when unwell and are a risk to others, especially as attitudes to and possibilities for working from home have developed. The pandemic has demonstrated that social and employment measures to ensure that people can afford not to go to work are a key element in protecting the vulnerable in society more widely. Normalising social distancing would require wider public health messaging and there is thus a distinction between measures aimed at the general population versus those targeted to people living with airways diseases. A substantial proportion of respondents thought that measures to reduce infection should apply to the general population as well as to themselves, especially during the ‘flu season. This should at least prompt public discussion about behaviours in enclosed public spaces where transmission risk is highest. Face coverings are probably most effective at reducing *transmission* of infection rather than protecting the wearer, so the protection for an individual wearing one may be limited. Interestingly around 60% of respondents expressed the view that face coverings should be required for all in indoor spaces and on public transport during the ‘flu season. Acceptability of interventions is dependent on many factors including cultural norms, such that it has already been commonplace in people from South East Asia to wear face coverings. In other jurisdictions the use of face coverings has become politicised which may further affect implementation, and some people with respiratory disease have been exempt from wearing face coverings because of a perceived effect on breathlessness which could also limit future adoption. The potential benefits of measures must be balanced against potential harms including costs and psychological harms such as stigma and drawing attention to disease status [24]. Acceptability of measures may change with time, particularly with changes in the impact of COVID-19 risk such as vaccination.

There are strengths and limitations to our work. Our survey was conducted online, so required a degree of digital literacy. The sample was large but a convenience sample and the extent to which respondents were representative of people with airways disease more broadly is unclear. The face validity of the responses is supported by the rational pattern of responses, with substantially greater support for measures that are likely to be effective such as greater support for the use of face coverings indoors compared to outdoors. Although the questions were framed to address the period “after COVID”, attitudes may change over time. However, even if only a small proportion of patients adhere to infection control measures, the simplicity and cost-effectiveness could justify highlighting their importance in guidelines and promoting implementation as part of exacerbation reduction strategies. Including such measures in guidelines may also prompt clinicians to encourage patients to adopt such interventions, thereby also increasing uptake. Furthermore, these non-pharmacological measures may appeal to people reluctant to comply with pharmacotherapy. Further work is required to optimise the behaviour-change interventions that would be necessary to maximise the benefit of continued respiratory virus infection control measures.

The impact of pandemic respiratory virus infection control measures indicate that local, national and international guideline committees in asthma, bronchiectasis and COPD should consider including advice on such interventions to reduce the burden of exacerbations. This is particularly important in the absence of a vaccine or drugs to combat rhinovirus – the commonest cause of acute exacerbations [5]. Our findings suggest that there is likely to be a significant level of acceptance by people with these conditions and infection control measures are affordable, not pathogen specific and mostly well tolerated [22]. Whilst no single measure had universal support, there was substantial support for a number of interventions enabling people living with chronic airways diseases to make informed choices about their own risk of exacerbation and how they choose to mitigate that risk. Recommendations to encourage the general public to take steps to protect vulnerable fellow citizens should also be supported by healthcare professionals.

## Data Availability

The data are the property of the Asthma UK - British Lung Foundation Partnership. Requests for data sharing will be considered.

## Acknowledgements

We thank all the survey respondents.

## References

1. Global Initiative for Asthma. Global Strategy for Asthma Management and Prevention 2020 Update. Available at https://ginasthma.org/wp-content/uploads/2020/04/GINA-2020-full-report_-final-_wms.pdf – last accessed March 22nd 2021.

2. Polverino E, Goeminne PC, McDonnell MJ, Aliberti S, Marshall SE, Loebinger MR, Murris M, Cantón R, Torres A, Dimakou K, De Soyza A, Hill AT, Haworth CS, Vendrell M, Ringshausen FC, Subotic D, Wilson R, Vilaró J, Stallberg B, Welte T, Rohde G, Blasi F, Elborn S, Almagro M, Timothy A, Ruddy T, Tonia T, Rigau D, Chalmers JD. European Respiratory Society guidelines for the management of adult bronchiectasis. Eur Respir J. 2017 Sep 9;50(3):1700629. doi: 10.1183/13993003.00629-2017. PMID: 28889110.

3. Global Initiative for Chronic Obstructive Lung Disease. Global Strategy for Prevention, Diagnosis and Management of COPD. Available at https://goldcopd.org/wp-content/uploads/2020/11/GOLD-REPORT-2021-v1.1-25Nov20_WMV.pdf. 2021 - last accessed March 22nd 2021.

4. Zhang Y, Morgan RL, Alonso-Coello P, Wiercioch W, Bała MM, Jaeschke RR, Styczeń K, Pardo-Hernandez H, Selva A, Ara Begum H, Morgano GP, Waligóra M, Agarwal A, Ventresca M, Strzebońska K, Wasylewski MT, Blanco-Silvente L, Kerth JL, Wang M, Zhang Y, Narsingam S, Fei Y, Guyatt G, Schünemann HJ. A systematic review of how patients value COPD outcomes. Eur Respir J. 2018 Jul 19;52(1):1800222. doi: 10.1183/13993003.00222-2018. PMID: 30002103.

5. Hewitt R, Farne H, Ritchie A, Luke E, Johnston SL, Mallia P. The role of viral infections in exacerbations of chronic obstructive pulmonary disease and asthma. Ther Adv Respir Dis. 2016 Apr;10(2):158–74. doi: 10.1177/1753465815618113. Epub 2015 Nov 26. PMID: 26611907; PMCID: PMC5933560.

6. Johnston SL, Pattemore PK, Sanderson G, Smith S, Lampe F, Josephs L, Symington P, O’Toole S, Myint SH, Tyrrell DA, et al. Community study of role of viral infections in exacerbations of asthma in 9-11 year old children. BMJ. 1995 May 13;310(6989):1225–9. doi: 10.1136/bmj.310.6989.1225. PMID: 7767192; PMCID: PMC2549614.

7. Nicholson KG, Kent J, Ireland DC. Respiratory viruses and exacerbations of asthma in adults. BMJ. 1993 Oct 16;307(6910):982–6. doi: 10.1136/bmj.307.6910.982. PMID: 8241910; PMCID: PMC1679193.

8. Bloom CI, Drake TM, Docherty AB, Lipworth BJ, Johnston SL, Nguyen-Van-Tam JS, Carson G, Dunning J, Harrison EM, Baillie JK, Semple MG, Cullinan P, Openshaw PJM; ISARIC investigators. Risk of adverse outcomes in patients with underlying respiratory conditions admitted to hospital with COVID-19: a national, multicentre prospective cohort study using the ISARIC WHO Clinical Characterisation Protocol UK. Lancet Respir Med. 2021 Mar 4:S2213-2600(21)00013-8. doi: 10.1016/S2213-2600(21)00013-8. Epub ahead of print. PMID: 33676593.

9. Gerayeli FV, Milne S, Cheung C, Li X, Yang CWT, Tam A, Choi LH, Bae A, Sin DD. COPD and the risk of poor outcomes in COVID-19: A systematic review and meta-analysis. EClinicalMedicine. 2021 Mar;33:100789. doi: 10.1016/j.eclinm.2021.100789. Epub 2021 Mar 18. PMID: 33758801; PMCID: PMC7971471.

10. UK National Health Service. Advice for people at high risk from coronavirus (clinically extremely vulnerable). Available at https://www.nhs.uk/conditions/coronavirus-covid-19/people-at-higher-risk/advice-for-people-at-high-risk - last accessed April 9th, 2021.

11. How COVID-19 is changing the cold and flu season. Nature 2020;588,388–390.

12. Berghaus TM, Karschnia P, Haberl S, Schwaiblmair M. Disproportionate decline in admissions for exacerbated COPD during the COVID-19 pandemic. Respir Med. 2020 Aug 14:106120. doi: 10.1016/j.rmed.2020.106120. Epub ahead of print. PMID: 32839072; PMCID: PMC7427552.

13. Chan KPF, Ma TF, Kwok WC, Leung JKC, Chiang KY, Ho JCM, Lam DCL, Tam TCC, Ip MSM, Ho PL. Significant reduction in hospital admissions for acute exacerbation of chronic obstructive pulmonary disease in Hong Kong during coronavirus disease 2019 pandemic. Respir Med. 2020 Sep;171:106085. doi: 10.1016/j.rmed.2020.106085. Epub 2020 Jul 12. PMID: 32917356; PMCID: PMC7354382.

14. Tan JY, Conceicao EP, Wee LE, Sim XYJ, Venkatachalam I. COVID-19 public health measures: a reduction in hospital admissions for COPD exacerbations. Thorax. 2020 Dec 3:thoraxjnl-2020–216083. doi: 10.1136/thoraxjnl-2020-216083. Epub ahead of print. PMID: 33273024; PMCID: PMC7716293.

15. McAuley H, Hadley K, Elneima O, Brightling CE, Evans RA, Steiner MC, Greening NJ. COPD in the time of COVID-19: an analysis of acute exacerbations and reported behavioural changes in patients with COPD. ERJ Open Res. 2021 Jan 18;7(1):00718–2020. doi: 10.1183/23120541.00718-2020. PMID: 33527075; PMCID: PMC7607968.

16. Sykes DL, Faruqi S, Holdsworth L, Crooks MG. Impact of COVID-19 on COPD and asthma admissions, and the pandemic from a patient’s perspective. ERJ Open Res. 2021 Feb 8;7(1):00822–2020. doi: 10.1183/23120541.00822-2020. PMID: 33575313; PMCID: PMC7734714.

17. Wee LE, Conceicao EP, Tan JY, Sim JXY, Venkatachalam I. Reduction in asthma admissions during the COVID-19 pandemic: consequence of public health measures in Singapore. Eur Respir J. 2021 Mar 2:2004493. doi: 10.1183/13993003.04493-2020. Epub ahead of print. PMID: 33653808; PMCID: PMC7926041.

18. Davies GA, Alsallakh MA, Sivakumaran S, Vasileiou E, Lyons RA, Robertson C, Sheikh A; EAVE II Collaborators. Impact of COVID-19 lockdown on emergency asthma admissions and deaths: national interrupted time series analyses for Scotland and Wales. Thorax. 2021 Mar 29:thoraxjnl-2020–216380. doi: 10.1136/thoraxjnl-2020-216380. Epub ahead of print. PMID: 33782079; PMCID: PMC8011424.

19. Huh K, Kim YE, Ji W, Kim DW, Lee EJ, Kim JH, Kang JM, Jung J. Decrease in hospital admissions for respiratory diseases during the COVID-19 pandemic: a nationwide claims study. Thorax. 2021 Mar 29:thoraxjnl-2020–216526. doi: 10.1136/thoraxjnl-2020-216526. Epub ahead of print. PMID: 33782081; PMCID: PMC8011422.

20. Shah SA, Quint JK, Nwaru BI, Sheikh A. Impact of COVID-19 national lockdown on asthma exacerbations: interrupted time-series analysis of English primary care data. Thorax. 2021 Mar 29:thoraxjnl-2020–216512. doi: 10.1136/thoraxjnl-2020-216512. Epub ahead of print. PMID: 33782080; PMCID: PMC8011425.

21. Cheng DO, Hurst JR. COVID-19 and ‘basal’ exacerbation frequency in COPD. Thorax. 2020 Dec 22:thoraxjnl-2020–216637. doi: 10.1136/thoraxjnl-2020-216637. Epub ahead of print. PMID: 33443208.

22. Jefferson T, Del Mar CB, Dooley L, Ferroni E, Al-Ansary LA, Bawazeer GA, van Driel ML, Nair S, Jones MA, Thorning S, Conly JM. Physical interventions to interrupt or reduce the spread of respiratory viruses. Cochrane Database Syst Rev. 2011 Jul 6;2011(7):CD006207. doi: 10.1002/14651858.CD006207.pub4. Update in: Cochrane Database Syst Rev. 2020 Nov 20;11:CD006207. PMID: 21735402; PMCID: PMC6993921.

23. Open Prescribing. Prednisolone. Available at https://openprescribing.net/chemical/0603020T0/ - last accessed 9th April 2021.

24. Philip K, Cumella A, Farrington-Douglas J, Laffan M, Hopkinson N. Respiratory patient experience of measures to reduce risk of COVID-19: findings from a descriptive cross-sectional UK wide survey. BMJ Open. 2020 Sep 9;10(9):e040951. doi: 10.1136/bmjopen-2020-040951. PMID: 32912958; PMCID: PMC7482474.

25. Philip KEJ, Lonergan B, Cumella A, Farrington-Douglas J, Laffan M, Hopkinson NS. COVID-19 related concerns of people with long-term respiratory conditions: a qualitative study. BMC Pulm Med. 2020 Dec 9;20(1):319. doi: 10.1186/s12890-020-01363-9. PMID: 33298023; PMCID: PMC7724437.

26. Halpin, D., Miravitlles, M., Metzdorf, N. and Celli, B., 2017. Impact and prevention of severe exacerbations of COPD: a review of the evidence. International Journal of Chronic Obstructive Pulmonary Disease, Volume 12, pp.2891–2908.

27. Little P, Stuart B, Hobbs F, Moore M, Barnett J, Popoola D et al. An internet-delivered handwashing intervention to modify influenza-like illness and respiratory infection transmission (PRIMIT): a primary care randomised trial. The Lancet. 2015;386(10004):1631–1639.

28. Wright L, Steptoe A, Fancourt D. Predictors of self-reported adherence to COVID-19 guidelines. A longitudinal observational study of 51,600 UK adults. The Lancet Regional Health – Europe 2021;4:100061

